# Effect of non-pharmaceutical interventions for containing the COVID-19 outbreak in China

**DOI:** 10.1101/2020.03.03.20029843

**Authors:** Shengjie Lai, Nick W Ruktanonchai, Liangcai Zhou, Olivia Prosper, Wei Luo, Jessica R Floyd, Amy Wesolowski, Mauricio Santillana, Chi Zhang, Xiangjun Du, Hongjie Yu, Andrew J Tatem

**Affiliations:** WorldPop, School of Geography and Environmental Science, University of Southampton, UK; Wuhan Center for Disease Control and Prevention, Wuhan, Hubei Province, China; Department of Mathematics, University of Tennessee, Knoxville, TN, USA; Computational Health Informatics Program, Boston Children’s Hospital, Boston, MA, USA; Department of Pediatrics, Harvard Medical School, Boston, MA, USA; Department of Epidemiology, Johns Hopkins Bloomberg School of Public Health, Baltimore, MD, USA; School of Public Health (Shenzhen), Sun Yat-sen University, Shenzhen, China; School of Public Health, Fudan University, Key Laboratory of Public Health Safety, Ministry of Education, Shanghai, China

**Keywords:** COVID-19, coronavirus, SARS-CoV-2, outbreak, non-pharmaceutical interventions, population movement, travel restriction, social distancing, model

## Abstract

**Background:** The COVID-19 outbreak containment strategies in China based on non-pharmaceutical interventions (NPIs) appear to be effective. Quantitative research is still needed however to assess the efficacy of different candidate NPIs and their timings to guide ongoing and future responses to epidemics of this emerging disease across the World.

**Methods:** We built a travel network-based susceptible-exposed-infectious-removed (SEIR) model to simulate the outbreak across cities in mainland China. We used epidemiological parameters estimated for the early stage of outbreak in Wuhan to parameterise the transmission before NPIs were implemented. To quantify the relative effect of various NPIs, daily changes of delay from illness onset to the first reported case in each county were used as a proxy for the improvement of case identification and isolation across the outbreak. Historical and near-real time human movement data, obtained from Baidu location-based service, were used to derive the intensity of travel restrictions and contact reductions across China. The model and outputs were validated using daily reported case numbers, with a series of sensitivity analyses conducted.

**Results:** We estimated that there were a total of 114,325 COVID-19 cases (interquartile range [IQR] 76,776 - 164,576) in mainland China as of February 29, 2020, and these were highly correlated (p<0.001, R^2^=0.86) with reported incidence. Without NPIs, the number of COVID-19 cases would likely have shown a 67-fold increase (IQR: 44 - 94), with the effectiveness of different interventions varying. The early detection and isolation of cases was estimated to prevent more infections than travel restrictions and contact reductions, but integrated NPIs would achieve the strongest and most rapid effect. If NPIs could have been conducted one week, two weeks, or three weeks earlier in China, cases could have been reduced by 66%, 86%, and 95%, respectively, together with significantly reducing the number of affected areas. However, if NPIs were conducted one week, two weeks, or three weeks later, the number of cases could have shown a 3-fold, 7-fold, and 18-fold increase across China, respectively. Results also suggest that the social distancing intervention should be continued for the next few months in China to prevent case numbers increasing again after travel restrictions were lifted on February 17, 2020.

**Conclusion:** The NPIs deployed in China appear to be effectively containing the COVID-19 outbreak, but the efficacy of the different interventions varied, with the early case detection and contact reduction being the most effective. Moreover, deploying the NPIs early is also important to prevent further spread. Early and integrated NPI strategies should be prepared, adopted and adjusted to minimize health, social and economic impacts in affected regions around the World.

**Funding:** Bill & Melinda Gates Foundation; EU Horizon 2020; National Natural Science Fund of China; Wellcome Trust.

**Research in context:** *Evidence before this study:* The COVID-19 outbreak has spread widely across China since December 2019, with many other countries affected. The containment strategy of integrated nonpharmaceutical interventions (NPIs) including travel bans and restrictions, contact reductions and social distancing, early case identification and isolation have been rapidly deloyed across China to contain the outbreak, and the combination of these interventions appears to be effective. We searched PubMed, Wanfang Data, and preprint archives for articles in English and Chinese published up to February 29, 2020, that contained information about the intervention of the COVID-19 outbreak. We found 15 studies that have investigated or discussed the potential effects of traveller screening, Wuhan’s lockdown, travel restrictions, and contact tracing in China or other countries. However, none of them comprehensively and quantitatively compared the effectiveness of various NPIs and their timings for containing the COVID-19 outbreak.

*Added value of this study:* To our knowledge, this is the most comprehensive study to date on quantifying the relative effect of different NPIs and their timings for COVID-19 outbreak containment, based on human movement and disease data. Our findings show that NPIs, inter-city travel restrictions, social distancing and contact reductions, as well as early case detection and isolations, have substantially reduced COVID-19 transmission across China, with the effectiveness of different interventions varying. The early detection and isolation of cases was estimated to prevent more infections than travel restrictions and contact reductions, but integrated NPIs would achieve the strongest and most rapid effect. Our findings contribute to improved understanding of integrated NPI measures on COVID-19 containment and can help in tailoring control strategies across contexts.

*Implications of all the available evidence:* Given that effective COVID-19-specific pharmaceutical interventions and vaccines are not expected to be available for months, NPIs are essential components of the public health response to the ongoing outbreaks. Considering the narrowing window of opportunity around the World, early and integrated NPI strategies should be prepared, deployed and adjusted to maximise the benefits of these interventions for containing COVID-19 spread.

## Introduction

As of February 28, 2020 the COVID-19 outbreak has caused 78,961 confirmed cases (2791 deaths) across China, with the majority seen in Wuhan City, and 4691 cases (67 deaths) reported in the other 51 countries.^1^ Further spread has occurred to all populated continents of the World, with many anticipating that a pandemic is approaching.^2,3^ As an emerging disease, effective pharmaceutical interventions are not expected to be available for months,^4^ and healthcare resources will be limited for treating all cases. Nonpharmaceutical interventions (NPIs) are therefore essential components of the public health response to outbreaks.^1,5-7^ These include isolating ill persons, contact tracing, quarantine of exposed persons, travel restrictions, school and workplace closures, and cancellation of mass gathering events.^5-7^ These containment measures aim to reduce transmission, thereby delaying the timing and reducing the size of the epidemic peak, buying time for preparations in the healthcare system, and enabling the potential for vaccines and drugs to be used later on.^5^ For example, social distancing measures have been effective in past influenza epidemics by curbing human-to-human transmission and reducing morbidity and mortality.^8-10^

Three major NPIs have been taken to mitigate the spread and reduce the outbreak size of COVID-19 across China.^11,12^ First, inter-city travel bans or restrictions have been taken to prevent further seeding the virus during the Chinese new year (CNY) holiday. People in China were estimated to make close to 3 billion trips over the 40-day CNY travel period from January 10 to February 18, 2020.^12,13^ A cordon sanitaire of Wuhan and surrounding cities in Hubei Province was put in place on January 23, 2020, just two days before CNY’s day on January 25. However, Wuhan’s lockdown is likely to have occurred during the latter stages of peak population numbers leaving the city before CNY, with around 5 million people likely leaving before the start of the travel ban, departing into neighbouring cities and other megacities in China.^14^ Since CNY’s day, travel restrictions in other provinces were also put in place across the country.

The second group of containment measures involves improving the screening, contact tracing, identification, diagnosis, isolation and reporting of suspected ill persons and confirmed cases.^11^ Since January 20, particularly in Wuhan, searches for cases, diagnosis and reporting have sped up across the country. Local governments across China encouraged and supported routine screening and quarantine of travellers from Hubei Province in an attempt to detect COVID-19 infections as early as possible. In Wuhan, where the largest number of infected people live, residents were required to measure and report ther temperature daily to confirm their onset, and those with mild and asymptomatic infections were also quarantined in “Fang Cang” hospitals, which are public spaces such as stadiums and conference centres that have been repurposed for medical care.^11^ The average interval from symptom onset to laboratory confirmation has dropped from 12 days in the early stages of the outbreak to 3 days in early February, highlighting how the efficiency of disease detection and diagnosis has greatly improved.^15,16^

Third, inner-city travel and contact restrictions were implemented to reduce the risk of community transmission. This involved limiting individual social contact, using personal hygiene and protective measures when people needed to move in public, and increasing the physical distance between those who have COVID-19 and those who do not.^11^ As part of these social distancing policies, Chinese government encouraged people to stay at home as much as possible, cancelled or postponed large public events and mass gatherings, and closed libraries, museums, and workplaces.^17,18^ Additionally, to fully cover the suspected incubation period of COVID-19 spread before Wuhan’s lockdown, the CNY and school holidays were also extended, with the holiday end date changed from January 30 to March 10 for Hubei province, and Feb 9 for many other provinces.^19-21^

The implementation of these NPIs has coincided with the rapid decline in the number of new cases across China, albeit at high economic and social costs.^15,16^ On February 17, the State Council required localities to formulate differentiated county-level measures for precise containment of the COVID-19 outbreak and the restoration of socioeconomy affected by the outbreak.^22^ The timing of implementing and lifting interventions is likely to have been and continue to be important, to take advantage of the window of opportunity to save lives and minimize the economic and social impact.^23,24^

The increasing numbers of cases of COVID-19 outside China and establishment of secondary transmission in multiple places highlights its pandemic potential. The best available scientific evidence is therefore required to design effective NPI strategies and disseminate this knowledge urgently to help policy makers assess the potential benefits and costs of NPIs to contain COVID-19 outbreaks. Some previous studies have preliminarily explored the lockdown of Wuhan,^25-27^ travel restrictions,^28-30^ airport screening,^31,32^ and the isolation of cases and contact tracing for containing virus transmission, respectively.^33,34^ The conclusions of these studies are persuasive, there are still key knowledge gaps on the effectiveness of different interventions.^15^ To fully justify the preparation, implementation, or cancellation of various NPIs, policy makers across the World need evidence as to the combination and timings of each, which remains lacking.

Based on near-real time human movement and disease data, here we conducted an observational and modelling study to develop a travel network-based modelling framework. We aimed to reconstruct COVID-19 spread across China and assess the effect of the three major groups of NPIs mentioned above. Given the expanding landscape of epidemics across the World, our findings contribute to improved understanding of the effect of NPI measures on COVID-19 containment and can help in tailoring control strategies across contexts.

## Methods

A travel network-based stochastic susceptible-exposed-infectious-removed (SEIR) model was built to simulate the COVID-19 spread between and within all prefecture-level cities in mainland China. Population movement data on human mobility across the country were used to estimate the intensity of travel restrictions and contact reductions. Data from illness onset to reporting of the first index case for each county were used to infer the changing timeliness of case identification and isolation across the course of the outbreak. The outputs of the model under NPIs were validated by using daily numbers of new cases reported across all regions in mainland China. Based on this modelling framework, the efficacy of applying or lifting non-pharmaceutical measures under various senarios and timings were tested and quantified.

### Data sources

Three population movement datasets, obtained from Baidu location-based services providing over 7 billion positioning requests per day,^35,36^ were used in this study to measure travel restrictions and social distancing across time and space. The first is an aggregated and de-identified dataset on near-real time daily relative outbound and inbound flow of mobile phone users for each prefecture-level city in 2020 (340 cities in mainland China were included) to understand mobility patterns during the outbreak. The daily outflow from each city since Wuhan’s lockdown and travel restrictions that were applied on January 23 were rescaled by the mean daily flow for each city during January 20 – 22 for comparing travel reductions across cities and years (Figure 1).

**Figure 1:**
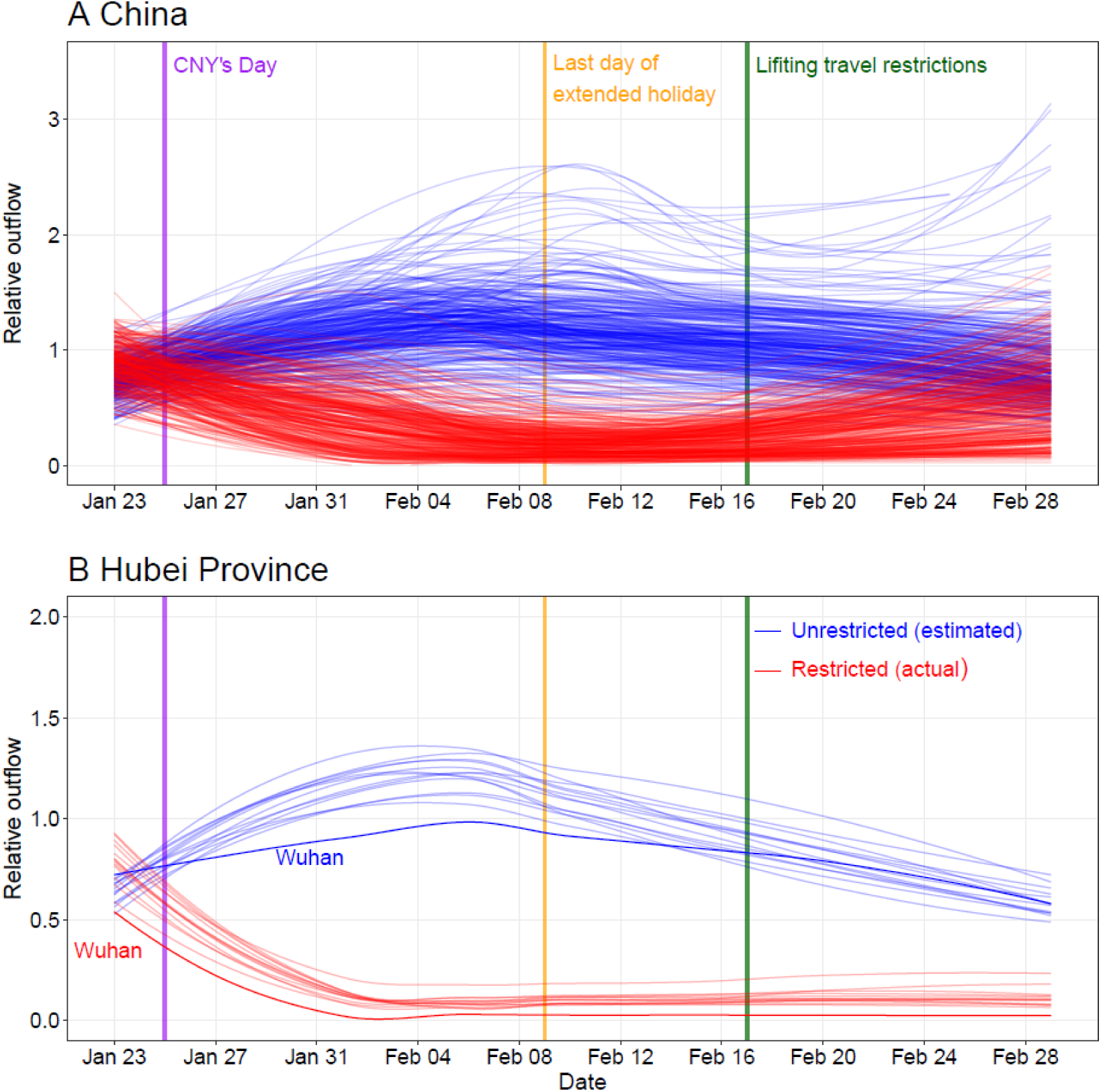
Relative daily volume of outbound travellers from cities (prefectural level) across mainland China during Chinese New Year (CNY) holiday, January 23^rd^ – February 29^th^, 2020. **(A)** All cities in mainland China. **(B)** Cities in Hubei province with Wuhan highlighted by using dark colours. Each blue line represents estimates of normal outflow by city under the scenario without travel restriction, following travel in previous years. The lines of relative volume were smoothed by using locally estimated scatterplot smoothing (LOESS) regression.

The second Baidu dataset is a historical relative movement matrix with daily total number of users at city level from December 26, 2014 to May 26, 2015, aligning with the 2020 CNY holiday period, for which the corresponding period is December 1, 2019 to April 30, 2020. We assumed that the pattern of population movements was the same in years when there were no outbreaks and interventions. Adjusted by the level of travel reductions derived from the 2020 dataset where applicable, the second dataset was used to simulate the COVID-19 spread and predict future transmission via population movements under various scenarios, with or without inter-city travel restrictions. Corresponding city-level population data in 2015 for modelling were obtained from the Chinese Bureau of Statistics.^37^

The third Baidu dataset measures daily population movements at county level (2862 counties in China) from January 26 through April 30, 2014, as described elsewhere.^38^ Based on the assumption that the pattern of population contact was consistent across years when there were no interventions, it was used to estimate inner-city travel and contact reduction under the outbreak and interventions. First, we aggregated data from county to city level and rescaled the daily flows since January 29, 2014 by the mean of the daily flow for the January 26 – 28 period, aligning with the date of Wuhan’s lockdown and the 2020 CNY holiday. Then, the rescaled first dataset for 2020 under interventions was compared with the 2014 dataset to derive the percentage of travel decline for each city. The percentages for cities were averaged by day to preliminarily quantify the intensity of contact reduction in China under NPIs (appendix Table S1), as the policies of travel restriction and social distancing measures were implemented and occurred at the same time across the country.

We also collated data of the first case reported by county across mainland China to measure the delay from illness to case report as a reference of the improved timeliness of case identification, isolation and reporting during the outbreak (appendix Table S2). The daily reported number of COVID-19 cases in Wuhan City, Hubei Province and other provinces were also used to futher validate our results. These case data were collated from the websites of national and local health authorities, news media, and publications (appendix note).^14,39,40^

### Data analysis

We constructed a travel network-based SEIR modelling framework (the code of model is available online at https://github.com/wpgp/BEARmod) for before-and-after comparable analyses on NPI efficacy. First, we simulated the COVID-19 spread across a metapopulation, where each population represented a city across China. Within each population, numbers of susceptible, exposed, infectious, and recovered/removed people were tracked per day.^3^ The epidemiological parameters estimated for the early stage of the outbreak in Wuhan were used to parameterise the epidemic before widely implementing the NPIs.^41^ During each timestep, infected people first recovered or were removed at an average rate *r*, where *r* was equal to the inverse of the average infectious period. We used the median of time lags from illness onset to reported case as a proxy of the average infectious period, indicating the improving case identification and isolation under improved interventions (appendix Table S2). Exposed people then became infectious at a rate *ε*, where *ε* was the inverse of the average time spent exposed but not infectious, based on the estimated incubation period (5.2 days, 95% confidence interval [CI] 4.1 - 7.0).^41^

The number of new people that could become exposed was calculated based on the daily contact rate *c* and the number of infectious people in the city *I*_*i*_, and this was turned into a number of newly exposed people after multiplying by the fraction of people in *i* who were susceptible (accounting for potential encounters with already-infected people, which did not lead to a new infection). The daily contact rate *c* was the basic reproduction rate (*R*_0_, 2.2, 95%CI 1.4 - 3.9) divided by the average days (5.8, 95%CI 4.3-7.5) from onset to first medical visit and isolation,^41^ then weighted by the level of daily contact (appendix Table S1). Finally, infectious people moved between cities, where the probability of moving from city *i* to city *j*(*p*_*ij*_) was equal to the proportion of smartphone users who went from city *i* to city *j* in the corresponding day from the Baidu dataset in 2015, accounting for the travel restrictions in 2020.

In this model, stochasticity occurred through variance in numbers of people becoming exposed, infectious, and removed/recovered, as well as variance in numbers of people moving from one city to another. Newly-infected people, recovered people, and numbers of people who moved were calculated for each city, each day, by drawing from *Poisson* distributions, where the probability of each person transitioning between states was *ε, r*, and *p*_*ij*_ respectively. By modelling the COVID-19 epidemic in this way, we could simulate the incidence of COVID-19 cases, accounting for variance in recovery, infection, and movement across many simulation runs (1000). Additionally, the incidence since the NPIs were implemented would be affected both by infections before and after interventions. Then, we could use this model to test the transmission of COVID-19 under various intervention scenarios and timings, as well as the potential of further transmission after the lifting of travel restrictions and contact distancing measures on 17 February 2020.

The estimates of the model for the outbreak under current NPIs as the baseline scenario were compared with reported COVID-19 cases across time and space. The sensitivity and specificity were also calculated to examine the performance of the model in predicting the occurrence of COVID-19 cases at city level across China. The relative effect of NPIs were quantitatively assessed by comparing estimates of cases under various NPIs and timings with that of the baseline scenario. We also conducted a series of sensitivity analyses to understand the impact of changing epidemiological parameters on the estimates and uncertainties of intervention efficacy. R version 3.6.1 (R Foundation for Statistical Computing, Vienna, Austria) was used to perform data collation and analyses.

### Ethical approval

Ethical clearance for collecting and using secondary data in this study was granted by the institutional review board of the University of Southampton (No. 48002). All data were supplied and analysed in an anonymous format, without access to personal identifying information.

## Results

As of February 29, 2020, a total of 79,824 COVID-19 cases were reported in mainland China, with most cases (61%) having occurred in Wuhan (Table 1). The outbreak increased exponentially prior to CNY (Figure 2). However, the peak of epidemics across the country quickly appeared through implementing strong and comprehensive NPIs, including dramatic reductions in travel and contact, and significant improvements in the timeliness of case detection and reporting across the country (Figure 1 and appendix Tables S1 and S2). We estimated that there were a total of 114,325 COVID-19 cases (interquatile range [IQR] 76,776 – 164,576) in mainland China as of February 29, 2020, with 85% of these in Hubei Province. The epidemics outside of Hubei province likely reached a low level (< 10 cases per day) in late February or early March, while cities in Hubei Province may need another two or three weeks to reach same level as other provinces. The estimated epidemics and peaks were consistent with patterns of reported data by onset date, with a high correlation (p<0.001, *R*^*2*^*=*0.86) found across regions (Figure 2). The sensitivity and specificity of our model were 91% (280/308) and 69% (22/32), respectively, to predict a city with or without COVID-19 cases as of February 29, 2020.

**Table 1.** Reports and estimates of the COVID-19 cases in mainland China, as of February 29, 2020.

| Interventions and timing | Wuhan City, Hubei Province | Other cities in Hubei Province | Other provinces | Mainland China |
| --- | --- | --- | --- | --- |
| **Under current non-pharmaceutical interventions (NPIs)** |  |  |  |  |
| No. of cases reported (%)* | 49,122 (62) | 17,785 (23) | 12,917 (16) | 79,824 (100) |
| Estimated no. of cases (%) | 78,910 (69) | 18,503 (16) | 16,912 (15) | 114,325 (100) |
| Interquartile range | 51,952 - 111,280 | 11,029 - 28,685 | 9,499 - 27,033 | 76,776 - 164,576 |
| Dates of estimated peak number of cases | Jan 25 - 27, 2020 | Jan 24 - 26, 2020 | Jan 24 - 26, 2020 | Jan 25 - 27, 2020 |
| **Percentage (%) of cases that could have been prevented at earlier interventions** |  |  |  |  |
| One week ahead | 61 (45 - 79) | 71 (55 - 86) | 78 (62 - 90) | 66 (50 - 82) |
| Two weeks ahead | 84 (78 - 89) | 90 (82 - 94) | 91 (84 - 95) | 86 (81 - 90) |
| Three weeks ahead | 94 (92 - 96) | 97 (95 - 99) | 98 (97 - 99) | 95 (93 - 97) |
| **Estimated relative no. of cases at later interventions\*\*** |  |  |  |  |
| One week delay | 2.4 (1.6 - 3.5) | 3.1 (1.8 - 4.6) | 3.3 (2 - 5.4) | 2.6 (1.8 - 3.8) |
| Two weeks delay | 5.8 (4.0 - 8.6) | 8.6 (5.3 - 12.8) | 9.4 (6.1 - 14.6) | 6.7 (4.6 - 10.0) |
| Three weeks delay | 15.1 (9 - 21.1) | 22.6 (13.5 - 33.9) | 27.9 (17.5 - 42.8) | 17.6 (11.2 - 25.5) |
| **Estimated relative no. of cases under various NPIs\*\*** |  |  |  |  |
| Without inter-city travel restriction | 1.0 (0.6 - 1.3) | 1.1 (0.7 - 1.7) | 1.1 (0.7 - 1.7) | 1.0 (0.6 - 1.4) |
| Without inner-city contact reduction | 2.5 (1.7 - 3.7) | 2.6 (1.5 - 4.2) | 2.4 (1.2 - 4.0) | 2.6 (1.7 - 3.7) |
| Without case early detection and isolation | 5.0 (3.3 - 6.9) | 5.6 (3.2 - 8.4) | 5.1 (2.5 - 8.4) | 5.0 (3.3 - 7.1) |
| Without all interventions above | 51.4 (33.2 - 71.2) | 91.6 (57.6 - 132.5) | 124.7 (77.4 - 180) | 67.3 (43.7 - 93.7) |

\* The reported data of COVID-19 cases were obtained from the Chinese National Health Commission as of February 29, 2020.
\*\* Referring to the median of estimates under current interventions and timing.
The median and interquartile range of estimates are provided here.

**Figure 2:**
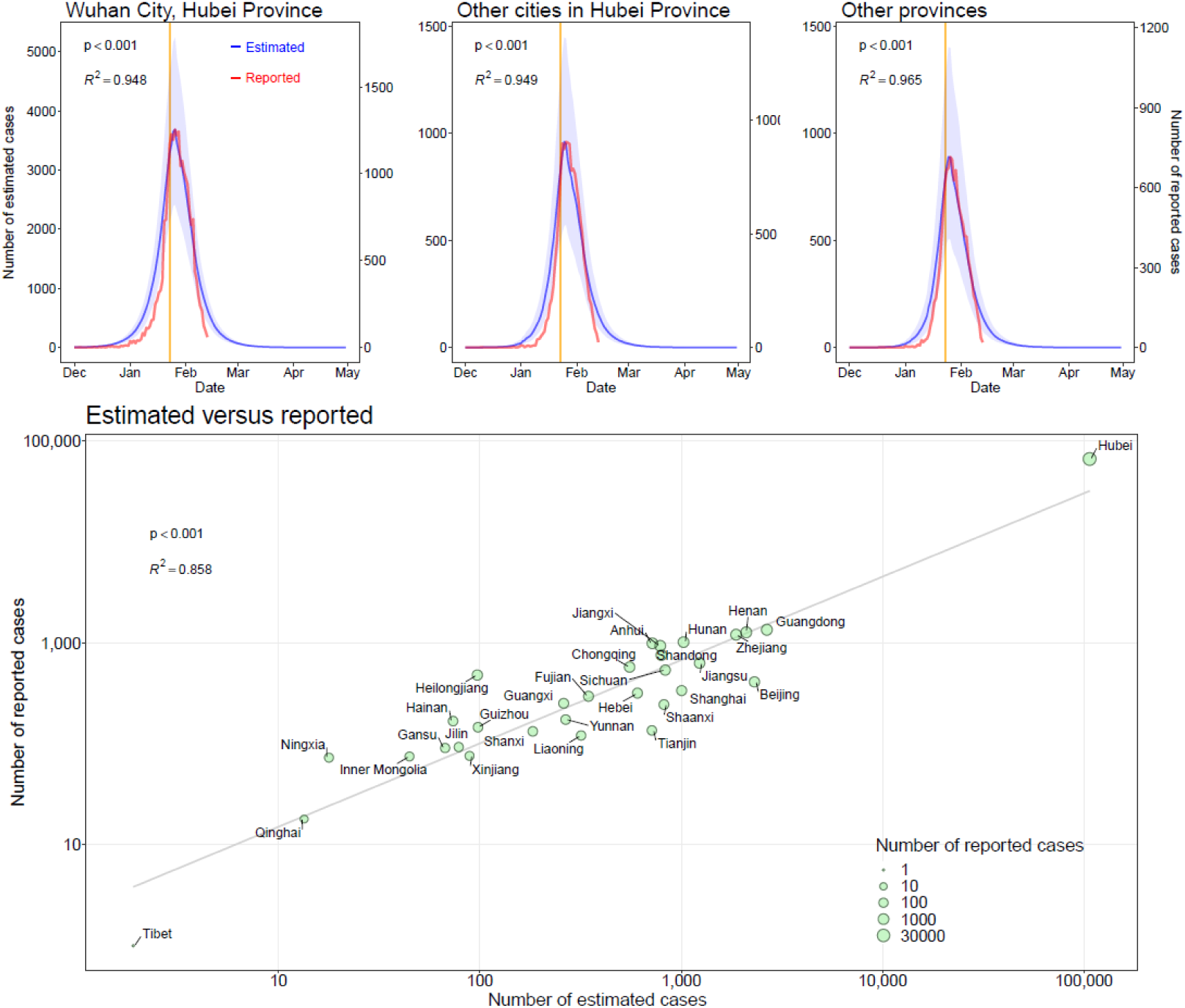
Estimated and reported epicurves of COVID-19 outbreak in China. Vertical lines: orange – date of Wuhan’s lockdown; purple – Chinese New Year’s day. The median and interquatile range (blue) of estimates of COVID-19 cases are presented with reported cases (red) by date of illness onset as of February 13, 2020. The reported data of COVID-19 cases in the scatterplot were obtained from the Chinese National Health Commission, as of February 29, 2020.

We found that without NPIs, the number of COVID-19 cases would increase rapidly across China, with a 51-fold increase in Wuhan, a 92-fold increase in other cities in Hubei, and 125-fold increase in other provinces, as of February 29 (Table 1). However, the apparent effectiveness of different interventions varied (Figure 3 and appendix Figure S1). The lockdown of Wuhan might not have prevented the seeding of the virus from the city, as the travel ban was put in place at the latter stages of outbound travel prior to CNY’s day (Figure 3). Nevertheless, if inter-city travel restrictions were not implemented, cities and provinces outside of Wuhan would have received more cases from Wuhan, and the affected geographic range would have expanded to the remote northern and western areas of China (Figure 4a and appendix Figure S2). Generally, the early detection and isolation of cases was estimated to quickly and substantially prevent more infections than contact reduction and social distancing across the country (5-fold versus 2.6-fold), but without the intervention of contact reductions, in the longer term, the epidemics would increase exponentially. Therefore, integrated NPIs would achieve the strongest and most rapid effect on COVID-19 outbreak containment (Table 1).

**Figure 3:**
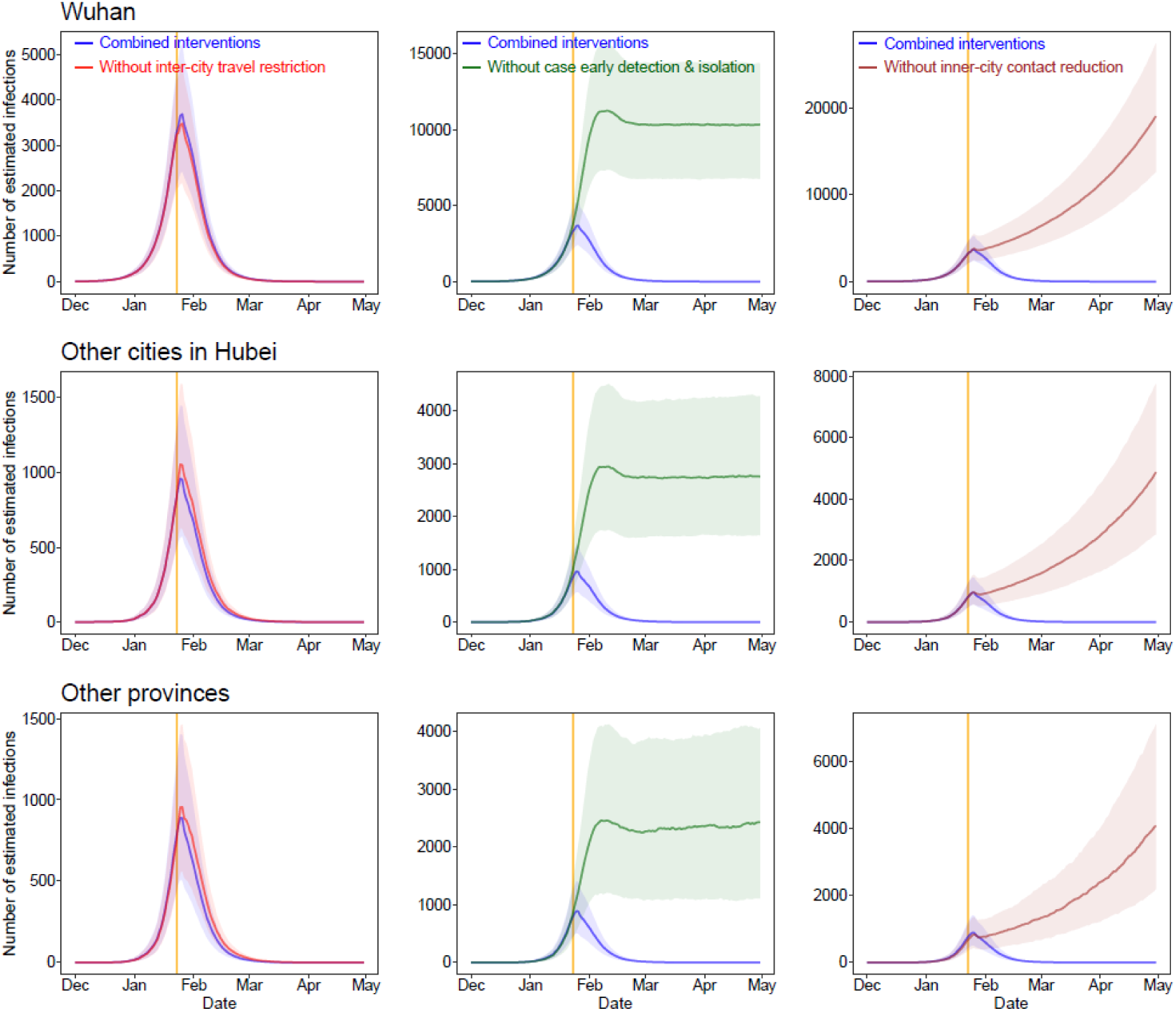
Estimated epicurves of COVID-19 outbreak under various scenarios with or without non-pharmaceutical interventions (NPIs) by region. The blue lines present estimated transmission under current NPIs, and each other line represents the scenario without one type of intervention. The median and interquartile range of estimates are provided here. The orange vertical line indicates the date of Wuhan’s lockdown on January 23, 2020.

**Figure 4:**
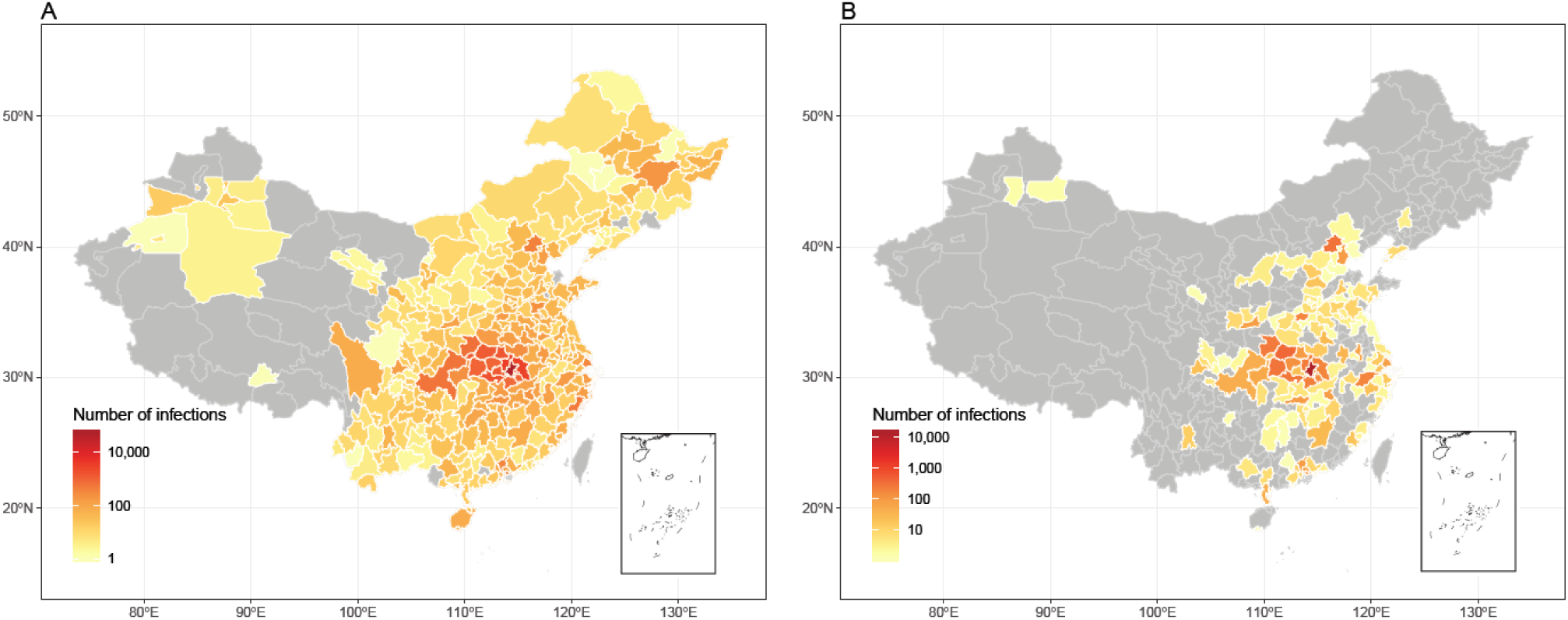
Affected areas of COVID-19 in mainland China as of February 29, 2020, under current interventions but with different timings. (A) Affected areas under interventions implemented at actual timing. A total of 308 cities reported cases, based on the data obtained from national and local health authorities, as of February 29, 2020. (B) Affected areas under interventions implemented at two weeks earlier than actual timing, with an estimate of 130 cities affected.

The timings of intervention implementation are also critical. The number of cases could be dramatically reduced by 66%, 86%, and 95%, respectively, if the NPIs could be conducted one week, two weeks, and three weeks earlier than the actual timing across the country (Figure 5). Moreover, the geographical range of affected areas would shrink from 308 cities to 192, 130, and 61 cities, respectively (Figure 3 and appendix Figures S3 and S4). Addtionally, if population contact resumed to the normal levels seen in previous years, the lifting of travel restrictions since February 17 might cause the epidemic to rise again (Figure 5 and appendix Figure S5). Therefore, the social distancing intervention should be continued for several months. Additionally, sensitivity analyses suggested that our model could have robustly measured relative changes of efficacy of various interventions on containing the COVID-19 outbreak under different epidemiological parameters and transmission senarios (appendix Figures S6-S12).

**Figure 5:**
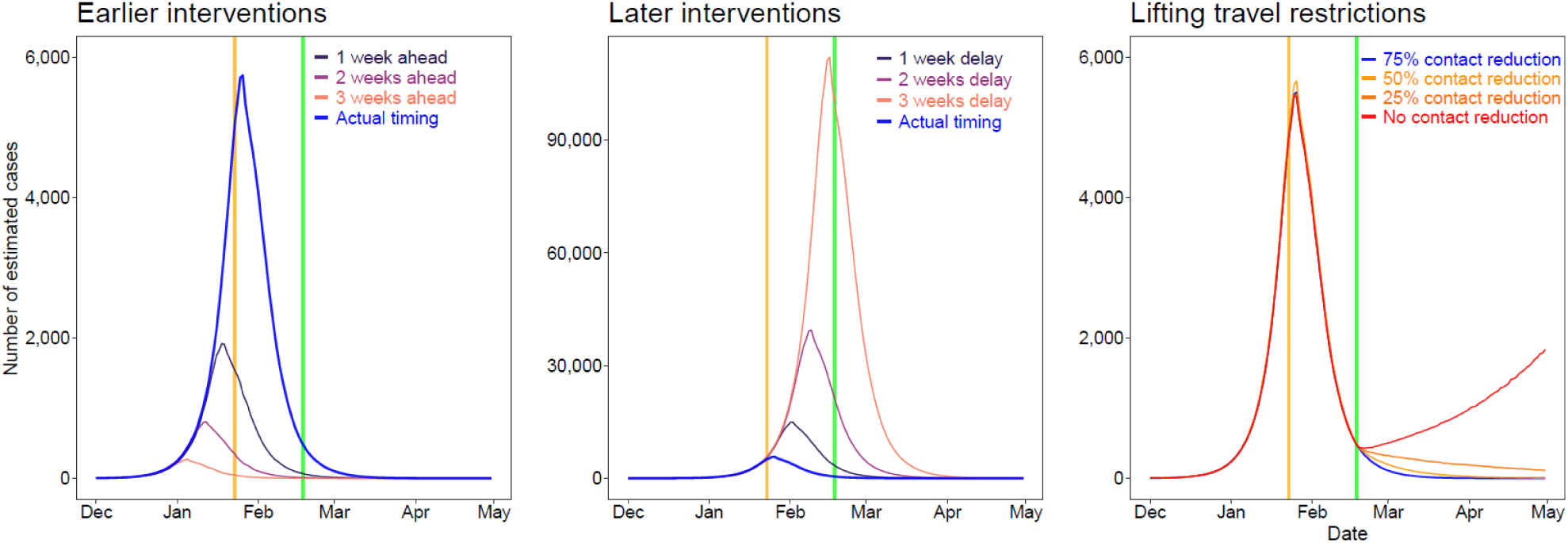
Estimates of the COVID-19 epidemic under various scenarios of intervention timing, travel restriction and contact reduction. Vertical lines: orange – date of Wuhan’s lockdown; purple - CNY’s Day; green – date of lifting of travel restrictions. The epidemics under various intervention timings were estimated under current non-pharmaceutical interventions. We estimated the COVID-19 spread under different population contact rates after lifting inter-city travel restrictions across the country on Feburary 17, 2020.

## Discussion

Our findings show that combined NPIs, inter-city travel restrictions, social distancing and contact reductions, as well as early case detection and isolations, have substantially reduced COVID-19 transmission across China. The lifting of inter-city travel restrictions since February 17, 2020, aiming to minimize the socioeconomic impact, does not appear to lead to an increase in cases if social distancing intervention can be maintained. Additionally, earlier implementation of interventions could have significantly reduced the magnitude and geographical range of the COVID-19 outbreak that has occurred in China. China’s vigorous, multifaceted response is likely to have prevented a far worse situation, which would have accelerated spread globally. The lessons drawn from China provide robust evidence and provide a preparation window and fighting chance for containing the spread of COVID-19 in other regions around the World.^15,16^

Three points raised by our findings are important. First, they support and validate the idea that population movement and close contact has a major role in the spread of COVID-19 within and beyond China,^3,14^ indicating the global risk of a pandemic via travellers infected with this virus. As the lockdown of Wuhan happened at the late stage of movement before CNY, travel restrictions did not halt the seeding of the virus from Wuhan, but it likely prevented extra cases being exported from Wuhan to a wider area. The second point is that the importance and effects of various NPIs differed. Compared to travel restrictions, improved detection and isolation of cases as well as the social distancing likely had a greater impact on the containment of outbreak. The social distancing intervention reduced contact with people who travelled from the epicentre of the epidemic, who were encouraged to quarantine at home. This is likely to have been especially helpful in curbing the spread of an emerging pathogen to the wider community, and reduced the spread risk from asymptomatic or mild infections.^5^ Third, given travel and work resuming in China, the country should consider at least partial continuation of NPIs to ensure that the COVID-19 outbreak is sustainably controlled. For example, early case identification and isolation should be maintained, and social distancing and personal hygiene are still proposed. Teleworking at home and staggered shifts are considered for mitigating COVID-19 transmission in workplaces or during the commute to and from work. Although the number of reported cases in the COVID-19 outbreak in China has decreased rapidly since February, we cannot be certain that the SARS-CoV-2 can be eradicated, as occurred for SARS-CoV.^42^

Ours is the most comprehensive study yet in which the effect of NPIs on COVID-19 transmission has been quantitatively assessed. Our model framework accounts for daily interactions of populations, interventions between and within cities, as well as the inherent statistical uncertainty associated with paucity of epidemiological parameters, before and after the interventions. Together with near-real time population movement and case data, our approach can be used for risk assessment for near real-time estimation of the effectiveness of different NPIs in the ongoing outbreaks in different countries. Our network-based SEIR model is methodologically robust and built on the basic SEIR models previously used to predict COVID-19 transmission in its early stages.^3^ Additionally, we assessed the effect of interventions by comparing estimates under various scenarios. Based on sensitivity analyses of multiple parameters, our results on the relative effects of NPIs are robust to the possibility of changes in parameters. Considering the delay in case reporting, our approach and findings can provide critical and early evidence for outbreak control decision-making.

However, our study has several limitations. First, as our simulations were based on the parameters estimated for the cases found in the early stage of the outbreak in Wuhan,, which might not account for the asymptomatic and mild infections, our study may underestimate the total number of infections. However, public awareness and enchanced case searching remained high throughout the study period, and a high proportion of infections was likely to have been detected, with nearly all reported cases eventually subjected to laboratory testing. Second, our findings could be affected by bias and confounding because the modelling is based on observations over a short period. Although we have shown that the apparent fall in incidence of COVID-19 since CNY’s day in China is likely to be attributed to the interventions taken, we cannot rule out the possibility that the decrease was caused by varying timings and intensities of various NPIs taken in different areas as well as some other unknown seasonal factors, e.g. temperature and absolute humidity.^43,44^ Third, our models and findings were based on some assumptions on parameterizations. If the epidemiological parameters of COVID-19 transmission in other cities across China differed with estimates from the outbreak at the early stage where no NPIs were in place in Wuhan, then our estimates of the effectiveness of interventions in reducing COVID-19 transmission could be biased. Although previous studies have supported the consistent seasonality of travel patterns across years in China and other countries,^14,36^ the magnitude and pattern could change year by year.

Additionally, inner-city travel restrictions and population contact reductions might not be highly correlated, and other data sources and further investigations are needed to explore this. Fourth, some coverage biases of mobile phone and Baidu users likely exist. Though a high percentage of the population owns mobile phones in China,^45^ the mobile user group still does not cover specific subgroups of the population, particularly children, and not all mobile owners use the Baidu location-based service. Therefore, our population movement data may provide an incomplete picture, and the spatiotemporal and demographic variations in the behaviour of phone users could have biased population distribution and travel estimates. Last, we only measured the main groups of NPIs and other interventions might also contributed to the outbreak containment, further research is needed to elaborate the effect of each intervention.

Because of the pandemic potential of the virus, should the outbreak spread widely in other countries, it will put a substantial burden on local health systems and society. From a public health standpoint, our results highlight that countries facing potential spread of COVID-19 should consider proactively planning NPIs and relevant resources for containment, given how the earlier implementation of NPIs could have lead to significant reductions in size of the outbreak in China. The results here provide some guidance for countries as to the likely effectiveness of different NPIs at different stages of an outbreak.

Suspected and confirmed cases should be identified, diagnosed, isolated and reported as early as possible to control the source of infection, and the implementation of cordon sanitaires or travel restrictions for significantly affected areas may prevent seeding the virus to wider regions. Reducing contact and increasing social distance, together with improved personal hygiene, e.g. hand washing, can protect vulnerable populations and mitigate COVID-19 spread at the community level, and these interventions should be promoted throughout the outbreak to avoid the resurgence. As called for by the World Health Organization, and backed up by our findings for China here, early and integrated NPI strategies should be prepared, deployed and adjusted to maximise benefits of these interventions and minimize health, social and economic impacts in affected regions, considering the narrowing window of opportunity around the World.^2,16^

## Data Availability

The data of COVID-19 cases reported by county, city, and province across China are availalable from data sources detailed in the Supplementary, and the average days from illness onset to report of the first case by each county used in the modelling are detailed in appendix Tabel S2. The mobile phone datasets analysed during the current study are not publicly available since this would compromise the agreement with the data provider, but the information on the process of requesting access to the data that support the findings of this study are available from the corresponding authors, and the data of travel and contact reductions derived from the datasets and used in our model are detailed in appendix Tabel S1.

## Contributors

SL designed the study, collected data, finalised the analysis, interpreted the findings, and wrote the manuscript. NWR built the model, analysed data, interpreted the findings, and wrote the manuscript. LZ, DW, and JX collected data, interpreted the findings, commented on and revised drafts of the manuscript. JRF, OP, and WL built the model, commented on and revised drafts of the manuscript. CZ collected data, interpreted the findings and commented on and revised drafts of the manuscript. AJT interpreted the findings and revised drafts of the manuscript. AW, XD, and HY interpreted the findings and commented on and revised drafts of the manuscript. All authors read and approved the final manuscript.

## Acknowledgments

We thank staff members at disease control institutions, hosptials, and health administractions across China where outbreaks occurred for field investigation, administration, and data collection. We thank Baidu Inc. sharing population movement data. We also thank Yanyan Zhu and Shuhao Lai for collating online data. This study was supported by the grants from the Bill & Melinda Gates Foundation (OPP1134076, OPP1195154); the European Union Horizon 2020 (MOOD 874850); the National Natural Science Fund of China (81773498); National Science and Technology Major Project of China (2016ZX10004222-009); Program of Shanghai Academic/Technology Research Leader (18XD1400300). AJT is supported by funding from the Bill & Melinda Gates Foundation (OPP1106427, OPP1032350, OPP1134076, OPP1094793), the Clinton Health Access Initiative, the UK Department for International Development (DFID) and the Wellcome Trust (106866/Z/15/Z, 204613/Z/16/Z). HY is supported by funding from the National Natural Science Fund for Distinguished Young Scholars of China (No. 81525023); Program of Shanghai Academic/Technology Research Leader (No. 18XD1400300); and the United States National Institutes of Health (Comprehensive International Program for Research on AIDS grant U19 AI51915).

## Data sharing

The data of COVID-19 cases reported by county, city, and province across China are availalable from data sources detailed in the Supplementary, and the average days from illness onset to report of the first case by each county used in the modelling are detailed in appendix Tabel S2. The mobile phone datasets analysed during the current study are not publicly available since this would compromise the agreement with the data provider, but the information on the process of requesting access to the data that support the findings of this study are available from Dr Shengjie Lai, and the data of travel and contact reductions derived from the datasets and used in our model are detailed in appendix Tabel S1.

